# Untargeted lipidomics of non-small cell lung carcinoma shows differentially abundant lipid classes in cancer vs non-cancer tissue

**DOI:** 10.1101/2021.03.16.21253733

**Authors:** Joshua M. Mitchell, Robert M. Flight, Andrew N. Lane, Hunter N.B. Moseley

## Abstract

Lung cancer is the leading cause of cancer death worldwide and non-small cell lung carcinoma (NSCLC) represents 85% of newly diagnosed lung cancers. The high mortality rate of lung cancer is due in part to the lack of effective treatment options for advanced disease. A major limitation in the development of effective treatment options is our incomplete understanding of NSCLC metabolism at a molecular level, especially lipids. Improvements in mass spectrometry combined with our untargeted assignment tool SMIRFE enable the systematic and less biased examination of NSCLC lipid metabolism.

Lipids were extracted from paired tumorous and non-tumorous lung tissue samples from 86 patients with suspected resectable stage I or IIa primary NSCLC and were analyzed using ultra-high resolution Fourier transform mass spectrometry. Pathological examination of the samples revealed that the majority of the samples were primary NSCLC; however, the disease group does include examples of metastases of other cancers and several granulomas. Information-content-informed (ICI) Kendall tau correlation analysis revealed correlation and co-occurrence patterns consistent with significant changes in lipid profiles between disease and non-disease samples. Lipids assigned using the program SMIRFE (Mitchell et al., 2019) were analyzed for differential abundance, followed by machine learning to classify the SMIRFE formula assignments into lipid categories. At the lipid category level, sterol abundances were consistently higher in diseased versus non-diseased lung tissues at statistically significant levels.

The statistically significant increase in sterol abundances in primary NSCLC compared with non-cancerous lung tissue suggests for treatment of primary NSCLC a possible therapeutic role for statins and nitrogenous bisphosphonates, pharmaceuticals that inhibit endogenous sterol biosynthesis. This hypothesis is consistent with previous epidemiological studies that have identified a therapeutic role for statins in the treatment of NSCLC but were unable to identify a molecular mechanism for this effect. Additionally, the majority of the consistently increased sterol abundances belong to the sterol ester subcategory, suggesting increased SCD1 and ACAT1 activity. SCD1 expression is a known negative prognostic indicator for survival in NSCLC. In our study, a large fraction of the NSCLC samples displayed this phenotype; however, SCD1 mutants would be unexpected in all of these samples. This suggests that this metabolic phenotype may be shared across multiple genetic subtypes of NSCLC. Thus, inhibitors of SCD1 and other enzymes involved in the production of this metabolic phenotype could have utility in the treatment of many genetic subtypes of NSCLC.

## Introduction

Lung cancer remains the most common cause of cancer death worldwide (Kanitkar et al., 2018) with approximately 85% of newly diagnosed lung cancers belonging to the non-small cell lung carcinoma subtype (Molina et al., 2008). The high mortality of lung cancer, NSCLC included, is partially explained by the insidious and silent nature of the progression of early-stage disease, but also the lack of effective therapeutic options for advanced disease. Although improvements have been made in the fields of NSCLC treatment, especially for adenocarcinomas with actionable mutations such as EGFR, ALK, certain K-RAS variants, and more recently with checkpoint inhibitors (Onoi et al., 2020), resistance often sets in (Liu et al., 2020) (Gettinger et al., 2017) (Walsh & Soo, 2020); and the overall 5-year survival rate remains low (<30%) (*SEER*Explorer: An interactive website for SEER cancer statistics*, 2021).

Treatment options for NSCLC vary with disease stage as well as genetic subtype. For low-stage disease, surgery remains the most common and most effective treatment option (Uramoto & Tanaka, 2014), especially when combined with adjuvant chemotherapeutic drugs (Betticher, 2005). For advanced disease, chemotherapy and/or radiotherapy (Sandler et al., 2000) (Pirker et al., 1995) (Wozniak et al., 1998) (Vansteenkiste et al., 2013) represents the primary first-line treatment option. Therapeutics targeting the epidermal growth factor receptor (EGFR) (Paez et al., 2004) (Shepherd et al., 2004), which is often mutated in NSCLC tumors in patients of East-Asian origin (Chang et al., 2006) and female never smokers, vascular endothelial growth factor (VEGF) (Piperdi et al., 2014) (Ferrara et al., 2005), anaplastic lymphoma kinase (ALK) (Crino et al., 2011) (Kim et al., 2014), and the PD-1/PD-L1 immune checkpoint (Sunshine & Taube, 2015) have increased the number of therapeutic options available for patients with advanced disease. However, while offering different side-effect profiles than traditional chemotherapy (Sgambato et al., 2016), overall survival of advanced NSCLC remains poor (Spigel et al., 2015) and many patients fail to express these particular biomarkers and are unsuitable for these therapies.

Finding drug targets for every genetic subtype of NSCLC would be very challenging and time-consuming. Alternatively, it is well known that cancer metabolism differs substantially from non-cancer metabolism due to metabolic reprogramming, a phenomenon that is essential to cancer development (Hanahan & Weinberg, 2011). As multiple genetic mutations may result in the same metabolic phenotypes, drugs targeting these metabolic processes may be efficacious for multiple genetic subtypes of NSCLC.

There are many examples of metabolic reprogramming in NSCLC (Sellers et al., 2015) (Hensley et al., 2016) (Sellers et al., 2019). In particular, isotope labeling studies in vivo have shown that both glycolysis and the TCA cycle are highly active in many NSCLC tumors, resulting in higher rates of glucose oxidation in NSCLC compared to surrounding non-NSCLC tissue (Hensley et al., 2016). Increased TCA activity provides the precursors needed for several anabolic processes needed for cell proliferation, but this requires concomitant increased anapleurosis, such as enhanced pyruvate carboxylation (Sellers et al., 2015), to maintain TCA cycling. Additionally many NSCLC tumors oxidize several other substrates with a preference for non-glucose substrates such as lactate (Faubert et al., 2017) and glutamine (Hassanein et al., 2013) (Mohamed et al., 2014) (Metallo et al., 2012) from the tumor microenvironment, especially at higher perfusion rates (Hensley et al., 2016).

Lipid metabolism is also commonly altered in NSCLC. Key enzymes for lipid metabolism such as ATP citrate lyase (ACLY) (Osugi et al., 2015), fatty acid synthase (FASN) (Uramoto et al., 1999), and stearoyl-CoA desaturase 1 (SCD-1) (Huang et al., 2016) can be differentially expressed in NSCLC compared to non-cancer tissue. Overexpression of these genes enables enhanced production of many lipid classes and is also correlated with poorer clinical outcomes and tumor aggressiveness (Wang et al., 2002) (Visca et al., 2004) (Noto et al., 2013) (Huang et al., 2016) (Csanadi et al., 2015). Additionally, previous studies have demonstrated an association between serum cholesterol and survival in patients with resectable NSCLC (Sok et al., 2009) (as well as other forms of cancer (Jamnagerwalla et al., 2018) (Kitahara et al., 2011)) and an association between membrane cholesterol content and EGFR signaling activity (Ringerike et al., 2002). Collectively, these findings suggest that altered lipid production plays a key role in the development and progression of NSCLC and that enzymes in these pathways are promising drug targets.

An alternative to the costly *de novo* development of novel therapeutics is the repurposing of existing pharmaceuticals for new indications. For example, previous epidemiological studies have suggested that patients prescribed statins, inhibitors of HMG-CoA reductase in the mevalonate pathway, have better overall survival rates for a variety of cancers including NSCLC (Hung et al., 2017), even at late stages (Lin et al., 2016) and with concurrent gefitinib treatment (Han et al., 2011). If this effect could be attributed to inhibition of endogenous sterol production, this suggests that statins or other inhibitors of the mevalonate pathway such as nitrogenous bisphosphonates (Tsoumpra et al., 2015) could be repurposed for the treatment of NSCLC. However, the mechanism underlying this effect remains unclear, as statins have multiple known off-target effects besides inhibition of sterol biosynthesis including anti-inflammation (Diomede et al., 2001), immunomodulation (Sadeghi et al., 2001), and angiogenesis inhibition (Weis et al., 2002), all of which may contribute to this possible survival benefit.

As these examples illustrate, an improved understanding of the metabolic differences between NSCLC and non-cancerous lung tissue represents a major first step in constructing more complete models of NSCLC progression and ultimately the development of more effective therapeutics. Advances in ultra-high resolution mass spectrometry, particularly Fourier-transform mass spectrometry (FT-MS), provide significant analytical improvements, including the ability to resolve distinct isotopologues, and the detection of lower abundance metabolites. These capabilities combined with our in-house data processing pipeline, artifact mitigation (Mitchell et al., 2017; Mitchell et al., 2018), and untargeted assignment method called Small Molecule Isotope Resolved Formula Enumeration (SMIRFE) (Mitchell et al., 2019; Moseley et al., 2018) enables the assignment of molecular formulas to spectral features observed in NSCLC-derived lipid extracts without bias due to the incompleteness of existing metabolic databases (Mitchell et al., 2014) (Schrimpe-Rutledge et al., 2016). These assignments can be classified into lipid category and class using our machine learning methods (Mitchell et al., 2020) to investigate changes that occur in lipid profiles at the lipid category level.

## Materials and Methods

### Description of Paired Human Suspected NSCLC and Non-Disease Tissue Samples

Eighty-six patients with suspected resectable stage I or IIa primary non-small cell lung cancer (NSCLC) and without diagnosed diabetes were recruited based on their surgical eligibility. The extent of resection was determined by the surgeon in accordance with clinical criteria. Many of the specimens were obtained from wedge resections, which minimizes surgery time while the other specimens were acquired in less than 5 minutes after the pulmonary vein was clamped. Both techniques minimize ischemia in the resected tissues. Immediately after resection, the tumor was transected and a section of cancerous tissue and surrounding non-cancer tissue at least 5 cm away from the tumor were immediately flash frozen in liquid nitrogen and stored at <80 °C. On-site pathologists classified the samples into cancer subtype and stage when possible and confirmed cancer-free margins on parallel tissue samples. All samples were collected under a University of Louisville approved Internal Review Board (IRB) protocol or University of Kentucky IRB protocol and written informed consent was obtained from all subjects prior to inclusion in the study (Sellers *et al*., 2015).

The frozen samples were pulverized under liquid nitrogen to <10 μm particles using a Spex freezer mill and extracted using a modified Folch method, as previously described (Fan, 2012) (Fan et al., 2016). The lipid fraction was supplemented with 1 mM butylated hydroxytoluene and then dried by vacuum centrifugation at room temperature. Samples for FT-MS analysis were redissolved in 200-500 μL chloroform/methanol (2:1) supplemented with 1 mM butylated hydroxytoluene. Reconstituted lipid samples were diluted in in isopropanol/methanol/chloroform 4/2/1 (v/v/v) with 20 mM ammonium formate (95 μL of solvent for 5 μL of sample) before direct infusion.

### Mass Spectrometry Analysis of Paired Samples

Ultra-high resolution (UHR) mass spectrometry was carried out on a Thermo Tribrid Fusion Orbitrap (Thermo Scientific, San Jose, CA, USA) interfaced to an Advion Triversa Nanomate (Advion Biosciences, Ithaca, NY, USA) nanoelectrospray source using the Advion “type A” chip, also from Advion, inc. (chip p/n HD_A_384). The nanospray conditions on the Advion Nanomate were as follows: sample volume in wells in 96 well plate – 50 µL, sample volume taken up by tip for analysis – 15 µL, delivery time – 16 minutes, gas pressure – 0.4 psi, voltage applied – 1.5 kV, polarity – positive, pre-piercing depth – 10 mm. The Orbitrap Fusion Mass Spectrometer method duration was 15 minutes, and the MS conditions during the first 7 minutes were as follows: scan type – MS, detector type – Orbitrap, resolution – 450,000, lock mass with internal calibrant turned on, scan range (*m/z*) – 150-1600, S-Lens RF Level (%) – 60, AGC Target – 1e5, maximum injection time (ms) – 100, microscans – 10, data type – profile, polarity – positive. For the next 8 minutes, the conditions were as follows for the MS/MS analysis: MS properties: detector type – Orbitrap, resolution – 120,000, scan range (*m/z*) – 150-1600, AGC Target – 2e5, maximum injection time (ms) – 100, microscans – 2, data type – profile, polarity – negative; monoisotopic precursor selection – applied, top 500 most intense peaks evaluated with minimum intensity of 5e3 counts; data dependent MS^n^ scan properties: MS^n^ level – 2, isolation mode – quadrupole, isolation window (*m/z*) – 1, activation type – HCD, HCD collision energy (%) – 25, collision gas – Nitrogen, detector – Orbitrap, scan range mode – auto *m/z* normal, Orbitrap resolution – 120,000, first mass (*m/z*) – 120, maximum injection time (ms) – 500, AGC target – 5e4, data type – profile, polarity – positive. The ion transfer tube temperature was 275°C. (Yang *et al*., 2017). Two Thermo Tribrid Fusion instruments were used to acquire spectra. 53 patients (102 spectra total) were acquired from Fusion 1 and 40 patients (77 spectra total) were acquired from Fusion 2. Three of the patients had only a cancer or non-cancer sample acquired. SMIRFE was used to generate assignments for all spectra.

### Molecular Formula Assignment

The SMIRFE algorithm (Mitchell et al., 2019) assigns characterized peaks from the peak characterization method in a truly untargeted manner without a database of expected molecular formulas corresponding to known metabolites (Mitchell et al., 2019). This method queries peaks in the spectrum against a near-exhaustive set of molecular formulas given a description of an elemental molecular formula search space and a mass tolerance based on the digital resolution of the spectrum. Assignments are validated by testing their observed intensity ratios against their theoretically predicted ratios based on natural abundance.

### Lipid Categorization of Assigned Formulas

Assigned molecular formulas were classified into one or more lipid categories using a family of Random Forest classifiers (Mitchell et al., 2020) trained on entries from the LipidMaps (Sud et al., 2006) and HMDB (Wishart et al., 2017) databases. Entries from LipidMaps represent examples of known lipids and their categories, while the non-lipid entries from the HMDB were used as examples of known non-lipid biological compounds.

### Mass Spectrometry Peak Characterization

For each spectrum, each assigned formula was assigned a score equal to one minus the expectation value of the assignment (S) or two times S if the formula was assigned to a lipid category. Lipid categories that were previously observed to be overclassified by our models (neutral and acidic glycosphingolipids) were excluded. Voting is then performed using these scores after removing not_lipid or not_classified entries, and either returning the maximally abundant category or ‘multiple’ when there are ties across one or more lipid categories.

To minimize false assignment, non_lipid and multiple lipids were not considered in the analysis. All lipid isotopologue intensities were normalized by dividing the isotopologue intensity by the median intensity of all the lipid peaks in the sample. Missing values were replaced with a threshold value that is ½ of the lower confidence interval of the distribution of all log-transformed intensities from that tissue. Sample-sample correlations were calculated using an information-content-informed (ICI) Kendall-tau correlation that considers missing values in the calculation of concordance and discordance of pairs (Flight & Moseley, 2020).

### Quality Control of Patient Samples

To determine possible outlier samples, each sample’s median ICI-Kendall-tau correlation to other samples with the same disease label, as well as the fraction of lipids that could be considered as a possible outlier were calculated for each sample (Gierliński et al., 2015). Samples with low median correlation outlier values (lower than the lower limit minus the 1.5 times the interquartile range, as defined in the boxplot.stats R function) and high outlier lipid fraction (higher than the higher limit plus 1.5 times the interquartile range, as defined in the boxplot.stats R function) were removed from further analysis.

### Consistently assigned spectral feature (corresponded peak) generation and Differential Abundance Analysis

Our SMIRFE assignment method assigns isotope-resolved molecular formulas (IMFs) to characterized peaks in each spectrum. Each IMF represents an isotopologue of a given elemental molecular formula (EMF) (e.g. ^13^C_1_^12^C_5_^1^H_12_^16^O_6_ is an IMF representing the m+^13^C_1_ isotopologue of EMF C_6_H_12_O_6_). To minimize the effects of misassignment, only consistently assigned spectral features (i.e. corresponded peaks) are used in differential abundance analysis. Corresponded peaks are identified using an in-house method we have named EMF voting which is described in supplemental materials. EMF voting identified 3485 total corresponded peaks across all 156 spectra with 884 corresponded peaks present in 25% or more of either the disease or non-disease group of samples.

Differential abundance analysis was then performed using both LIMMA (Ritchie et al., 2015) (Phipson et al., 2016) and SDAMS (Li et al., 2019) (Li Y, 2020) on the normalized features (see above). No pairing of samples was considered for the differential analysis, given the large size of the dataset and the desire to detect differences that are distinct even across patient biological variance. An adjusted p-value cutoff of 0.001 was used to determine metabolite significance. All lipid IMFs identified by either LIMMA or SDAMS were considered in this analysis. P-values were adjusted using the Benjamini-Hochberg multiple testing adjustment (Thissen et al., 2002). Cancer samples were clustered via hierarchical clustering on the sample-to-sample distances calculated as one minus the ICI-Kendall-tau correlation. Cluster leaves were ordered using the dendsort package (Sakai et al., 2014). All data manipulations and statistical testing were carried out in R v 4.0.0. (Ihaka & Gentleman, 1996).

LIMMA identified 287 differentially abundant corresponded peaks while SDAMS identified 426 abundant corresponded peaks. Together LIMMA and SDAMS identified 426 total unique differentially abundant corresponded peaks with 287 corresponded peaks identified by both methods. Of the 426 total unique differentially abundant corresponded peaks, 268 of them were assigned to one lipid category and the rest either had multiple lipid categories, were classified as “not lipid”, or a classification was not generated by the software. Only the 268 with a single lipid category were analyzed further. The log2 fold changes for each categorized corresponded peak between disease and non-disease was then calculated.

### Lipid Category Enrichment of Significant Peaks

Using all of the peaks with a single assigned lipid category, hypergeometric enrichment of the significant peaks in each category was tested. This used the assigned lipid category as an annotation of the peak, and all of singly categorized lipids as the peak universe (541 peaks). The significantly more-abundant and less-abundant features (based on log2 fold-changes of cancer / non-cancer) were tested independently (up, 120; down, 148). Hypergeometric enrichment was performed using the categoryCompare2 R package (v 0.99.158) (Flight et al., 2014).

## Results

### MS Data Processing, Assignment Ambiguity and Quality Control of Samples

As previously described (Mitchell et al., 2019), our SMIRFE methodology reliably produces highly accurate molecular formula assignments but not necessarily unique assignments, especially at higher m/z where the molecular formula search space grows exponentially. Assignment ambiguity is further compounded by biological and analytical variance resulting from variance between biological units (in our case human patients), sample preparation, and limitations in dynamic range of the instruments, which can introduce high amounts of data sparsity across the dataset.

EMF voting identifies consistently assigned (i.e. corresponded) peaks and filtering these features to those present in at least 25% of samples for a given sample class limits the level of data sparsity. This subsequent reduction in data sparsity allows for the meaningful use of more traditional statistical approaches such as PCA that do not handle high data sparsity well. Even so, the high data sparsity present in the dataset significantly contributes to the low percent variance observed in the top principal components.

Additionally, our quality control measures allow us to detect outlier spectra that could represent failed sample preparation, poor injection, or non-primary or non-NSCLC tumors. Initially 179 patient samples were included in the analysis. After removal of 10 samples from unrelated secondary metastatic tumors and of 13 outlier samples based on quality control, 156 samples remained for differential abundance analysis.

### PCA Shows Separation of Disease and Non-Disease Samples

Principal component analysis (PCA), performed using the normalized intensity of the corresponded peaks, revealed a clear, if imperfect, boundary between disease and non-disease samples along principal component 2 (PC2, Figure 1). This indicates that a significant portion of the variance observed in the second principal component represents biological variance between the two classes of samples. Correlation heatmaps between the samples further supports this claim (Figure 2). Samples from non-cancerous lung tissue cluster well together with high correlation observed between most of these samples. Correlation within the disease class were less strong but still differentiated disease samples from normal. Further investigation of the correlation among the cancer samples reveals two groups of cancer samples with the larger group comprising the majority of the adenocarcinoma samples (Supplemental Figure 1). In both groups, samples were removed if their quality control scores were outliers in two different quality control metrics. These samples likely have data quality or spectral acquisition issues.

**Figure 1.**
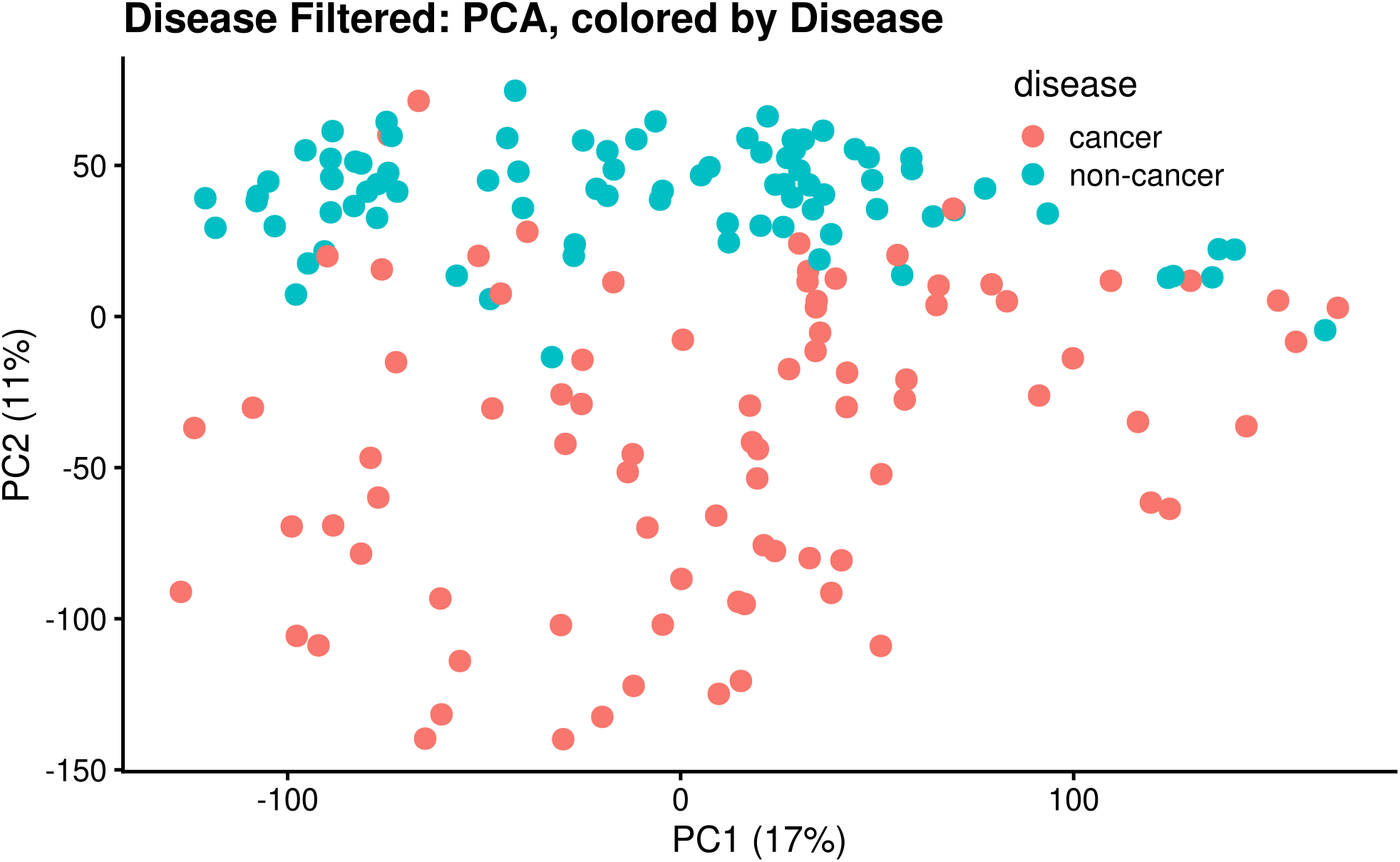
PCA by Disease. PCA was performed on the normalized intensities of the corresponded peaks present in at least 25% of a sample class and shows a clear separation between disease and non-disease that is most obvious along PC2. This separation is not present along PC1. The grouping of samples by disease class in PCA space clearly indicates that the corresponded peak intensities capture some of the biological variance between disease classes.

**Figure 2:**
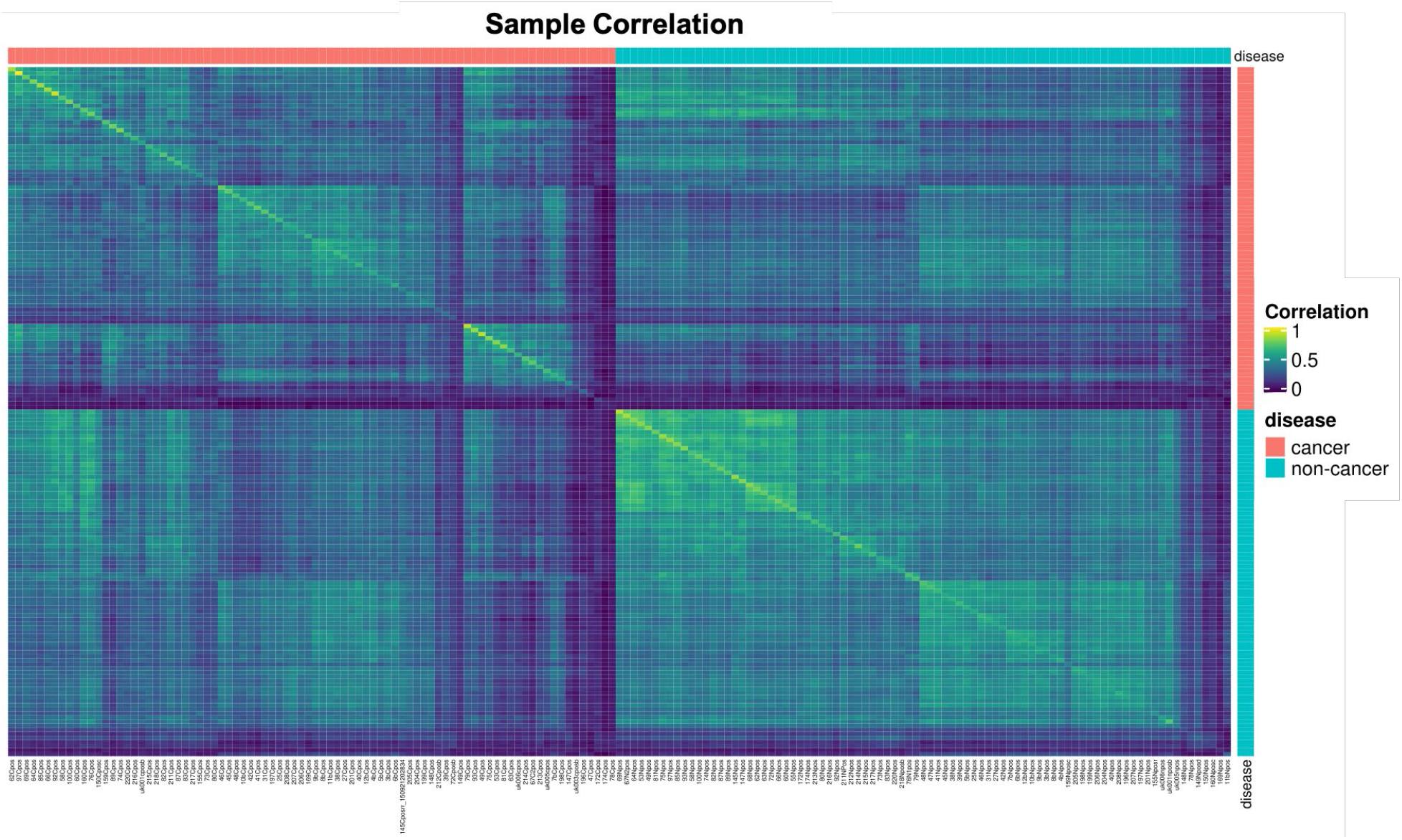
Correlation Heatmap by Disease. High correlation is observed between most non-disease samples and other non-disease samples and there are two subgroups with higher correlation observed. Correlation among cancer samples is less strong than the non-disease samples and there are multiple subgroups of samples that correlate more strongly with one another. Overall, the correlation between cancer samples is not as strong as the correlation between non-disease samples. In both disease classes, a smaller number of samples have poor correlation with samples from either class.

### Differential Abundance of Lipid Categories between disease and non-disease lung tissue

Lipids from five categories were observed: Fatty Acyls [FA], Glycerophospholipids [GP], Prenol Lipids [PR], Sphingolipids [SP] and Sterols [ST] across all samples. Of these five categories, only fatty acyls and prenols had no differentially abundant assignments. Additionally, few assignments were made to the fatty acyl or prenol categories. This may reflect the abundance of these lipids in our samples or their poor ionization in positive mode. Sphingolipids and sterols were significantly higher at the category level, whereas glycerophospholipids were significantly less abundant at the category level (Table 1).

**Table 1:** Differential Abundance Analysis Results. For each category of lipids, the number of observed more- and less-abundant features was recorded and compared to the number of expected more- and less-abundant features to statistically evaluate the differential abundance of that specific lipid category. P-values were calculated using a hypergeometric test and adjusted for multiple testing using the Benjamini-Hochberg technique (Benjamini & Hochberg, 1995). This revealed two statistically significant, more-abundant lipid categories and one statistically significant, less-abundant lipid category in cancer compared to non-cancer.

| Category | Number Observed | More Abundant Features |  |  | Less Abundant Features |  |  |
| --- | --- | --- | --- | --- | --- | --- | --- |
|  |  | Expected | Observed | p-adjust | Expected | Observed | p-adjust |
| Fatty Acyls [FA] | 12 | 2.662 | 0 | 1 | 3.283 | 0 | 1 |
| Glycerophospholipids [GP] | 210 | 46.58 | 32 | 1 | 57.449 | 71 | **0.0255** |
| Prenol Lipids [PR] | 5 | 1.109 | 0 | 1 | 1.368 | 0 | 1 |
| Sphingolipids [SP] | 291 | 64.547 | 75 | **0.04764** | 79.608 | 74 | 1 |
| Sterol Lipids [ST] | 23 | 5.102 | 13 | **0.00131** | 6.292 | 3 | 1 |

The patterns in fold changes are shown with respect to *m/z* in Figure 3. In cases where an IMF is observed in disease but not in non-disease, the fold-change is very high due to imputation of missing values. In the case of sphingolipids, more and less-abundant sub-populations appear to correlate with m/z, with cancer having higher concentrations of higher m/z sphingolipids on average. Sterols and glycerophospholipids show no such pattern with respect to m/z. Querying the top 5 most abundant unique EMFs of the more-abundant sterol IMFs against PubChem indicates that these assignments likely correspond to known sterol esters. Supporting this hypothesis is the observation that 12 of the 13 sterol EMFs assigned in our study were classified into the ST01 class of sterols from LipidMaps, which includes sterol esters.

**Figure 3.**
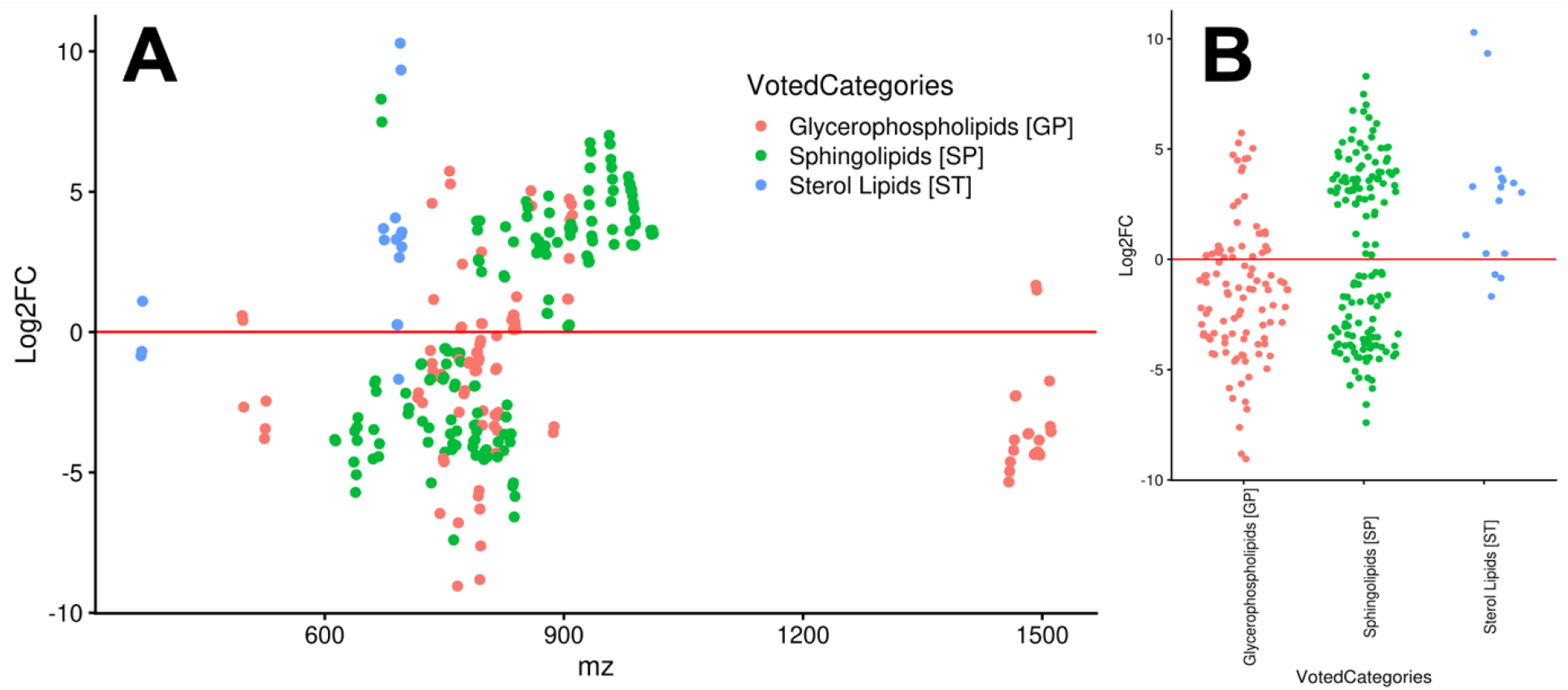
Log2 Fold Change by Category and m/z. The Log2 fold-change of consistently assigned, classified, and significantly changed lipids are shown in panel A with respect to m/z and by class in panel B. The extremely high fold-changes observed for some members of the sphingolipid and sterol lipid categories are due, in part, to imputed values. Most of the differentially abundant lipids occur in the 600 to 1000 m/z range; however, this region also has the highest density of assignments. Although no lipid category is exclusively more abundant, sterols are predominantly more abundant while substantial numbers of both sphingolipids and glycerophospholipids are more and less abundant.

### Lipid Category Correlation and Co-Occurence Heatmaps

Kendall tau correlation values between features were calculated using samples with non-zero corresponded peak normalized intensities for both features (Figure 4). At the lipid category level, we see distinct groups of correlated lipids within each category and in general, intra-category correlation is stronger than inter-category correlation. Strong intra-category correlations are expected, if regulation of these lipids is controlled at the category level and if the corresponded peaks are consistently and accurately assigned to lipid categories, which is why multi-classified corresponded peaks were dropped from these analyses.

**Figure 4.**
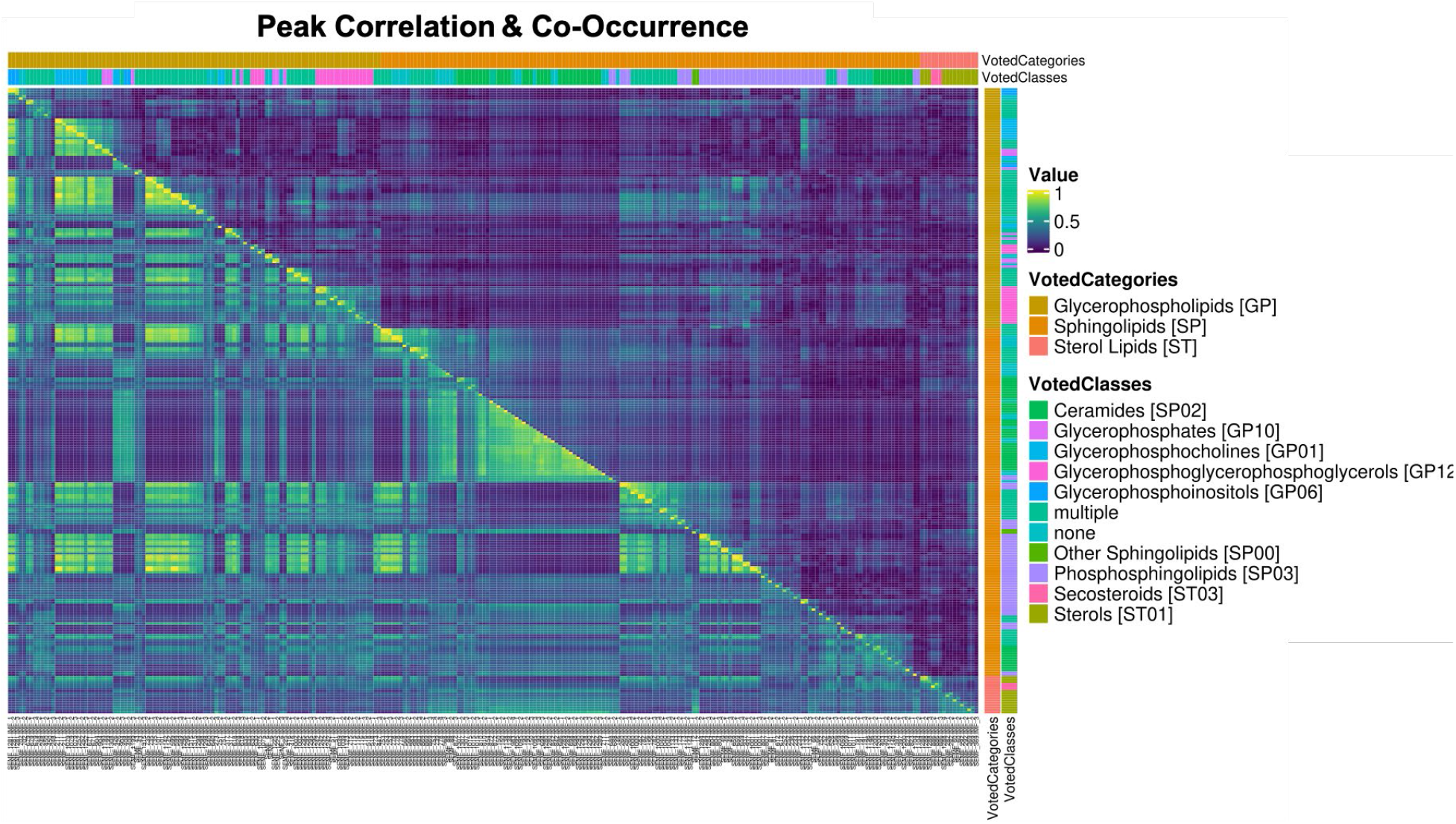
Peak Correlation and Peak Co-Occurrence Combined Heatmap. The bottom-left corner of Figure 4 shows the Kendall tau correlation between corresponded peak normalized intensities grouped by lipid category. Obvious strong correlations are seen between many members of the same lipid category. Additionally, some examples of inter-category correlation can be observed, suggesting possible co-regulation. Shown in the upper-right corner is the co-occurrence of lipids by lipid category and class. Notably there is a subset of sterols that co-occur with a sub-population of sphingolipids and a smaller sub-population of glycerophospholipids. The biological relevance of this co-occurrence is unclear. It is also possible that this co-occurrence simply reflects that these lipids were generally of lower relative abundance, which could artificially create the co-occurrence pattern.

Of the three groups, the glycerophospholipids collectively show the strongest intra-category correlations; however, some subgroups of glycerophospholipids are correlated with subgroups of sphingolipids and sterols. Within the sphingolipid category, there are distinct sub-populations of sphingolipids with a varying amount of intra-group correlation. Several groups correlate with the sterols as well and these groups of sphingolipids are not strongly correlated with other groups of sphingolipids. Finally, the sterols show the weakest intra-category correlation of the three categories, but there are two distinct subgroups of sterols correlated with one another. These findings suggest possible coregulatory mechanisms between these groups of lipids.

Patterns observed in the co-occurrence heatmaps of the lipids by class and category revealed more interesting findings. There are two sets of co-occurring sphingolipids which mirrors the findings from the correlation analysis. The sterols co-occur with one sub-population of sphingolipids; however, it is not the sub-population of sphingolipids that were correlated with the sterols.

### Reproducibility across Instruments and Clinical Environments

In this study, two sets of samples were analyzed using two different Thermo Tribrid Fusion instruments (FSN10115 and FSN10352, referred to as Fusion 1 and Fusion 2 respectively). Samples from Fusion 1 were derived only from patients undergoing resections at the University of Louisville, while Fusion 2 samples were derived from patients undergoing resections at both the University of Kentucky and the University of Louisville. PCA on the consistently assigned spectral features demonstrates that instrument and patient population specific variances are partially mitigated by our analysis as evidenced by the partial overlap between samples analyzed by the two instruments (Figure 5). The same analysis demonstrates a more obvious separation between cancer and non-cancer samples specifically along PC2 (Figure 1). These results argue for the reproducibility and consistency of our results and that the observed changes in lipid differential abundance more accurately reflect variance due to disease state and not on one of these potential confounding factors.

**Figure 5:**
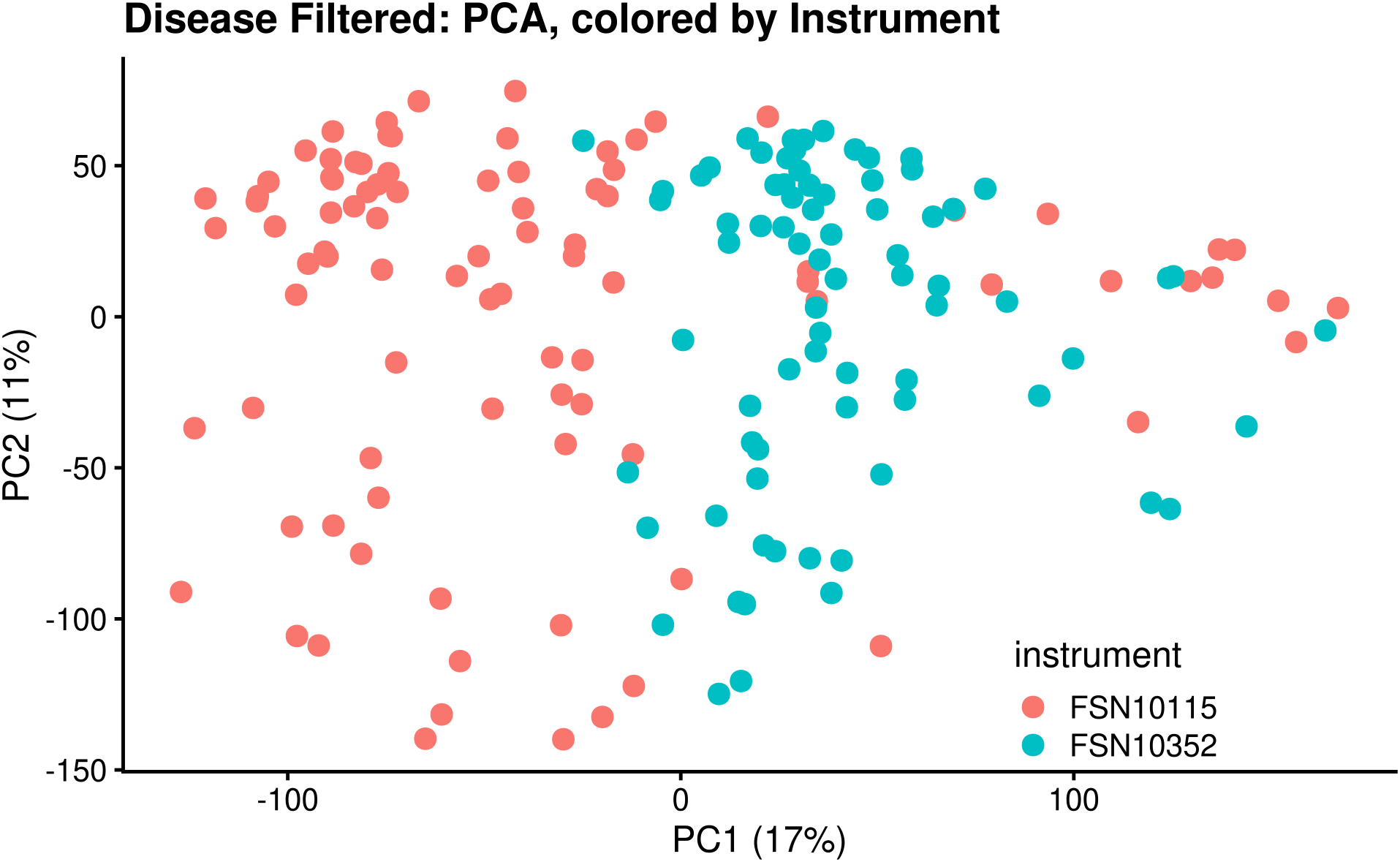
PCA of Consistently Assigned Spectral Features by Instrument. PCA on the consistently assigned spectral features colored by instrument demonstrates no clear decision boundary or primary component that groups the samples by instrument. A clearer decision boundary exists in the case of disease along PC2 as shown in Figure 1. This implies that features identified by SMIRFE assignment, EMF voting, and 25% group inclusion filtering represents cancer vs. non-cancer status rather than which instrument was used to perform the analysis or which patient cohort the sample was collected from.

## Discussion

### Sample Correlation Analysis Shows Evidence of Metabolic Reprogramming in NSCLC

Although metabolic reprogramming is ubiquitous in cancer, the number of distinct dysregulations of metabolic programs that can lead to a pro-cancer state is likely large even among a cohort of patients with the same type of cancer. We hypothesize that the lipid profiles of non-disease samples are expected to be more similar to one another than the lipid profiles of disease samples from the same patients. This expected high inter-tumor metabolic variance is analogous to previously observed high inter-tumor variance in gene expression (Hu et al., 2006), RNA editing (Paz-Yaacov et al., 2015), DNA methylation (Hansen et al., 2011), and mitochondrial DNA content (Mizumachi et al., 2008)

Likewise, this hypothesis is supported by the correlation patterns observed in Figure 2. Although individual genetic, metabolic, and environmental variance leads to some differences between lipid profiles of non-disease samples, the majority of the non-disease samples show higher correlation to one another with respect to the normalized intensities of shared corresponded peaks as compared to the disease samples. Additionally, the number of common corresponded peaks was higher in non-disease than in disease samples, arguing that in general, the non-disease samples have more similar lipid profiles to one another. The disease samples on the other hand displayed weaker correlation to one another, consistent with our hypothesis, with several distinct clusters of correlation observed. Further analysis of these clusters did not show a correlation with NSCLC subtype. The limited amount of correlation observed between disease and non-disease samples is explained by the sizable effect of metabolic reprogramming on some components of cellular metabolism.

Furthermore, these correlation patterns demonstrate that our assignment methods and data analysis pipeline, including our quality control measures, are successfully reducing assignment ambiguity and providing consistent assignments across samples as well as instrument and clinical environment. Random incorrect assignments would not result in the observed correlation patterns between samples nor the stronger correlations observed among peaks classified to the same lipid category.

### Lipid Category Correlation and Co-Occurrence

In addition to the differences in abundance at a lipid category level, our study also examined the correlation of normalized lipid intensity across samples as well as the co-occurrence of those features across samples. We observed strong intra-category correlation among most glycerophospholipids, among several sub-populations of sphingolipids and weak intra-category correlation with the sterols but two distinct sub-populations. Additionally, we observed inter-category correlation among some glycerophospholipids, sphingolipids, and one sub-population of sterols.

The *de novo* synthesis of lipids is complex and we hypothesize that these correlation patterns are the result of co-regulation across lipid categories possibly in combination with regulation at a lipid category level. A possible mechanism for this co-regulation is through steroid response element binding proteins (SREBPs), which are involved in regulating key enzymes in the sterol and glycerolipid biosynthetic pathways (Ericsson et al., 1997). Both SREBP-1 and SREBP-2 have been implicated in the control of lipid biosynthesis (Wen et al., 2018) and previous studies have shown that altered SREBP-1 signaling due to B7-H3 (aka CD276, an immune checkpoint protein) overexpression is correlated with aggressive NSCLC and increased glycerolipid production (Luo et al., 2017). However, the weaker correlation patterns observed among sphingolipids and glycerophospholipids could imply more incorrect assignments for these lipid categories, which is expected for sphingolipids due to our method’s tendency to overpredict that lipid category (Mitchell et al., 2020). This weaker correlation could also represent more complex regulatory mechanisms or a mixture of scavenging and *de novo* synthesis. Additionally, some correlation could be the result of multiple lipid categories sharing various precursor metabolites.

Correlation, however, is only part of the picture. Although many of the lipid categories were correlated to other members of their same category or class, not all members of each class are observed to co-occur with one another. Two distinct populations of sterols are clear in Figure 4B, one large and one small population of sterols that co-occur with one another. Additionally, each population of sterols co-occurs with other lipids from the other categories, implying that there are two distinct lipid profiles observed across the samples. The origin and clinical implications of these two lipid profiles is unclear, but one population could correspond to lung cancer that may respond to statins.

### Potential Clinical Implications

Our differential abundance analysis identified clear differences in the relative abundances of lipids between NSCLC tissue and non-cancerous tissue samples derived from the same patient. Notably, the relative abundances of a subset of sterols (Figure 4) were significantly and consistently higher in the disease samples compared to control. Although interfering with any of the metabolic pathways implicated by these findings could potentially have therapeutic roles in the treatment of NSCLC, sterol metabolism is the easiest to target with existing commonly-prescribed pharmaceuticals. As discussed earlier, increased sterol production is observed across multiple cancers and as such, this finding is not unexpected. While our results do not directly support a claim that statins have an anti-cancer effect through the inhibition of the mevalonate pathway, increased sterol production in cancer vs. non-cancer is a prerequisite for such a hypothesis to be true and our results are indirectly supportive of this hypothesis. More importantly, our methods provide a potential avenue to explore the effects of statins and other pharmaceuticals on lipid profiles in an untargeted and comprehensive manner.

The mechanisms resulting in the observed lipid profile changes from our study are not addressed here, and should be the object of future studies. Increased sterol and glycerophospholipids concentrations are consistent with direct (constitutively active mutants) or indirect (downstream signaling) activation of: i) EGFR, which promotes glycerophospholipids and sterol biosynthesis (Sukhanova et al., 2013) (Gabitova et al., 2013); ii) SCD, which is required for some sterol esters (Bené et al., 2001) and glycerophospholipids (Balgoma et al., 2019); and iii) ACLY, which is essential for lipid biosynthesis (Zaidi et al., 2012). As mentioned previously, SREBPs might also be involved in these lipid profile changes (Luo et al., 2017) (Ericsson et al., 1997). Furthermore, if the presence of more-abundant sterol esters can be verified, the enzyme ACAT-1 becomes a promising target in NSCLC. Previous studies on pancreatic cancer have shown that inhibition of ACAT-1 prevents the conversion of cholesterol to sterol esters, which results in apoptosis due to elevated endoplasmic reticulum stress (Li et al., 2016).

One possible hypothesis explaining the observed differences is that acquisition of a high concentration sterol and glycerophospholipid metabolic phenotype may be necessary for the development of NSCLC. Therefore, multiple distinct genetic lesions may result in this metabolic phenotype. Thus, pharmaceutical intervention that directly targets the sterol component of this phenotype may be beneficial for a variety of genetically distinct NSCLC subtypes. Alternatively, these lipid profile differences may be a byproduct of other disease processes and not directly contributory to the development of disease. In this case, these metabolic changes would still be useful as biomarkers for genetic subtypes of NSCLC.

## Conclusions

The use of untargeted assignment tools such as SMIRFE combined with EMF voting and machine learning models for the prediction of lipid categories enables comprehensive lipid profiling in human derived samples. The accuracy of the resulting molecular formula assignment and lipid classification for paired disease and non-disease tissue samples from human patients predominantly with primary NSCLC is confirmed through observed patterns of stronger within-patient correlation and within-lipid category and class correlations. A subsequent differential abundance analysis identified a consistent and significant difference in lipid profiles between disease samples and controls. Most notably, sterols are significantly and consistently observed in higher relative concentrations in disease tissue than in neighboring non-disease tissue. These findings are consistent with known genetic lesions observed in NSCLC and with common metabolic alterations observed in cancer metabolic reprogramming.

Furthermore, the change in sterol profile between disease and non-disease has the potential to be clinically translatable. Statins and other mevalonate pathway altering drugs, which are known to improve outcomes in NSCLC patients, could be altering these lipid profiles as part of their mechanism of action. Although further studies are necessary to test this hypothesis, the ability to detect and quantify lipid profiles in an untargeted manner provides the capability to directly observe the metabolic effect of pharmacological interventions targeting the mevalonate, with respect to NSCLC lipid profiles. Which genetic markers correlate to our observed molecular phenotype remains unclear, but future genomic or transcriptomics datasets from a similar cohort of patients combined with our lipid profiling approach could identify the genetic markers for this metabolic phenotype. The possibility that multiple genetic lesions could result in the same or similar metabolic phenotype implies that anti-mevalonate pathway therapies could have a direct chemotherapeutic or adjuvant role in the treatment of many genetically distinct subtypes of NSCLC.

## Data Availability

All data and results presented here are available in a FigShare repository: DOI: 10.6084/m9.figshare.14199521 . This includes characterized peaks derived from Fourier transform mass spectra and all downstream analyses of these characterized peak lists.

https://doi.org/10.6084/m9.figshare.14199521

## Acknowledgements

The work was supported in part by grants NSF 1419282 (PI Moseley), NSF 2020026 (PI Moseley), NIH 1P01CA163223-01A1 (PD Lane), 1U24DK097215-01A1 (PD Higashi), and P30 CA177558 (B.M. Evers, PI). We thank Timothy Fahrenholz for his efforts in mass spectrometry data collection and Teresa Fan and Rick Higashi for their efforts in human tissue sample collection.

## Data Availability

All data and results presented here are available in a FigShare repository: DOI: 10.6084/m9.figshare.14199521. This includes characterized peaks derived from Fourier transform mass spectra and all downstream analyses of these characterized peak lists.

## Abbreviations

NSCLC: Non-Small Cell Lung Carcinoma
SMIRFE: Small Molecule Isotope Resolved Formula Enumeration
SCD-1: Stearoyl-CoA Desaturase-1
ACAT-1: Acetyl-CoA Acetyltransferase
EGFR: Epidermal Growth Factor Receptor
ALK: Anaplastic Receptor Tyrosine Kinase Gene
PD-1: Programmed cell Death protein 1
PD-L1: Programmed Death-Ligand 1
HMDB: Human Metabolite Database
ICI: Information-Content-Informed
IMF: Isotope-resolved Molecular Formula
EMF: Elemental Molecular Formula
PCA: Principal Component Analysis

## Supplemental Materials

### EMF Voting Description and IMF-Level Data Extraction

Elemental molecular formula (EMF) voting was used to match peaks across samples and determine the most likely assignments from SMIRFE. First, for each sample, the assignments are extracted, filtered, and scored. Those assignments containing sulfur are removed, and any assignments that contain only peaks that were marked as being questionable removed. Scores are calculated as 1 – E-value, and EMFs that were classified as lipids had their score multiplied by 2.

For each sample, peaks were grouped to a sample specific EMF by determining the list of shared EMFs across a group of peaks (grouped_EMF). Scores for each formula in the grouped_EMF in the sample were taken as the best score for that formula from available scores in the group. After all sample grouped_EMFs are generated, additional scores for a formula are considered in actual voting by looking for the same unadducted base EMF with different adducts and adding these scores into the final total score.

pseudo_EMFs are collections of grouped_EMFs across samples. They are generated across samples by iteratively merging grouped_EMFs with shared formulas, creating a new list of formulas in an EMF, and merging any pseudo_EMFs that have shared formulas again.

For each pseudo_EMFs, the most likely formulas are determined by voting. Voting uses the sum of EMF scores across samples, including those from the same formula with a different adduct. Those formulas with a total score in the top 90% of all total scores were considered “winning” formulas and kept as the “voted formula” set. Any peaks that did not originally have a “voted formula” were then checked to see if the peak location was within previously defined tolerance, and if the ordered peak intensities for the set of peaks from a sample are in the same order as the natural abundance probabilities of IMFs for the voted EMF. If so, then the peaks are kept as having the “voted formula” set.

After voting, all of the peaks are checked to make sure that the peak locations are within previously defined m/z specific cutoffs. Any peak outside of it’s specific cutoff is removed.

Finally, those pseudo_EMFs that share greater than 50% of their peaks are merged together, and voting on the EMFs is performed again.

Peak locations from our custom data processing pipeline are reported both in m/z and frequency, which are derived from the m/z values. The frequency values are more reliable (i.e., more consistent with less variance), but differ between instruments. Therefore, for each set of samples from a particular instrument, we use high confidence assignments (e-value <= 0.1, m/z <= 600) to derive a frequency cutoff using the mode of the distribution of frequency standard deviations across both groups of samples. This frequency cutoff is used to do EMF voting within an instrument. The m/z standard deviations of the voted peaks are then fit to m/z using a generalized additive model, and the standard deviation of the predicted m/z values are multiplied by 2 to derive an m/z cutoff so that voting can be performed across the instruments.

Patient Info Table

demographic_info_NSCLC.xls

**Supplemental Figure 1:**
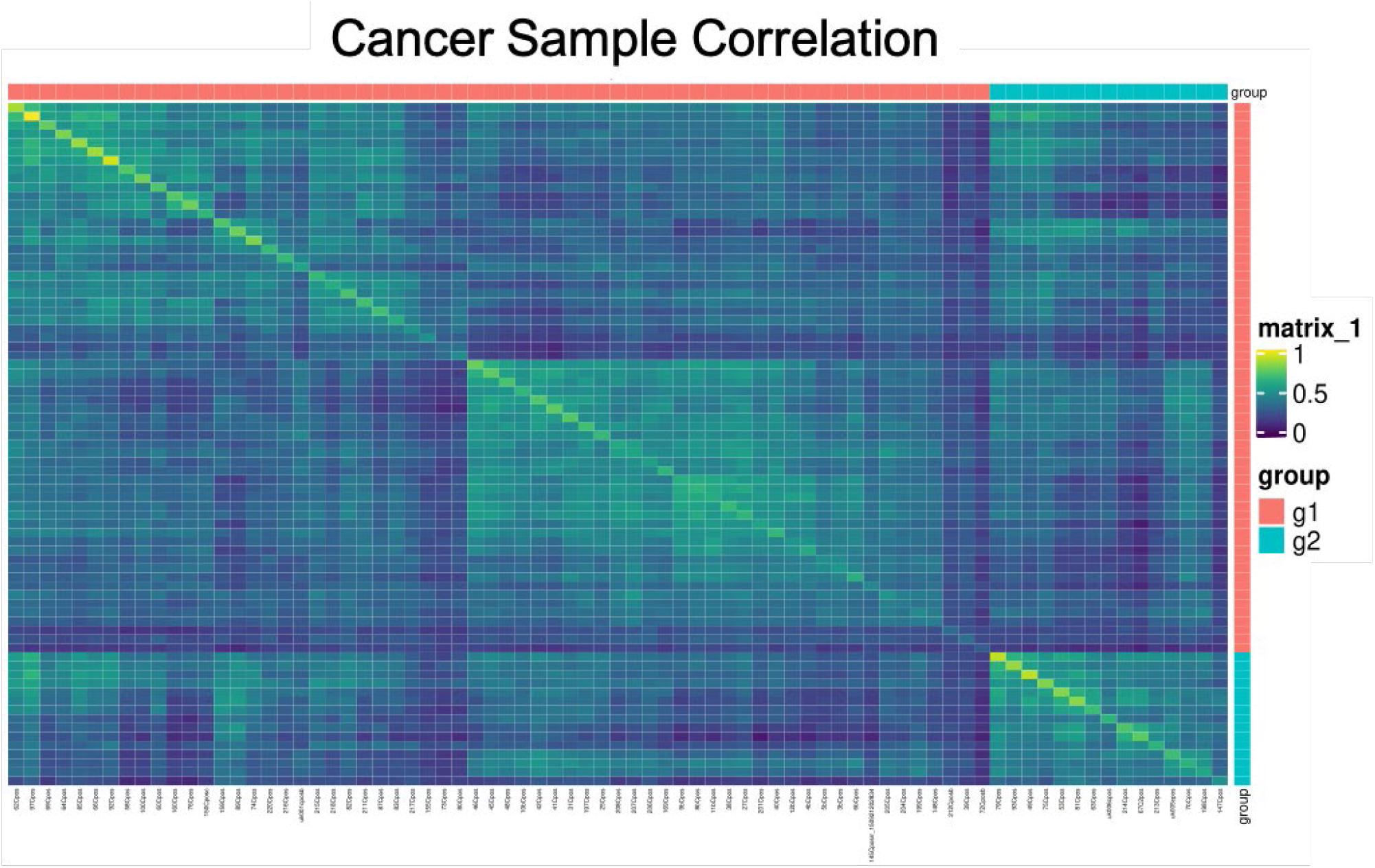
Further analysis of correlation observed between cancer samples reveals two distinct groups of cancer samples. Group 1 comprises 22 of the 26 adenocarcinoma samples and Group 2 the remaining 4. Group 1 appears to comprise at least two subgroups of cancer samples; however, neither of these subgroups represents one subtype of NSCLC.

